# Beneficial effects of a standardised saffron extract (Safr’Inside^TM^) on premenstrual syndrome: a multicentric, double-blind, randomized, placebo-controlled trial

**DOI:** 10.64898/2026.10.19.26365879

**Authors:** Camille Pouchieu, Roberta Villa, Fabio Amone, Line Pourtau, Benjamin Moras, Vincenzo Nobile

## Abstract

**Background:** Premenstrual syndrome (PMS) affects nearly half of women of reproductive age and can substantially impair quality of life, daily functioning, and psychological well-being. Although saffron extract has been proposed as a potential non-pharmacological approach for PMS management, clinical evidence remains limited. This randomized, multicentric, double-blind, placebo-controlled trial evaluated the efficacy and tolerability of saffron extract in reducing PMS symptoms.

**Methods:** A total of 110 healthy women aged 18–45 years with mild-to-moderate PMS were randomized to receive saffron extract (30 mg/day; Safr’Inside^TM^) or placebo for three menstrual cycles. The primary endpoint was the Premenstrual Assessment Form–Short Form (PAF-SF) total score. Secondary endpoints included PAF-SF affect, pain, and water retention subscales, and the Patient Health Questionnaire-4 (PHQ-4) total score. Efficacy analyses were performed in the intention-to-treat (ITT) and per-protocol populations using linear mixed-effects models.

**Results:** Saffron supplementation resulted in a significantly greater reduction in overall PMS symptoms than the placebo (difference in slope for PAF-SF total score: -1.88; 95% confidence interval [CI] -2.64 to -1.12; p<0.001). Between-group differences in change from baseline were significant at 7 days (-2.42; 95% CI -4.79 to -0.04; p=0.046), 35 days (-3.40; 95% CI -5.78 to -1.02; p=0.010), and 63 days of supplementation (-5.96; 95% CI -8.34 to -3.58; p<0.001). Significant improvements were also observed for affective symptoms (p=0.003), pain (p<0.001), water retention (p<0.001) from the PAF-SF, and psychological distress as assessed by the PHQ-4 total score (p=0.011). No adverse events were reported.

**Conclusions:** Daily supplementation with saffron extract over three menstrual cycles significantly improved overall PMS symptom severity compared with placebo in women with mild-to-moderate PMS. Benefits were also observed for affective, pain, and water retention symptoms. Saffron extract was well-tolerated and may represent a promising non-pharmacological option for women experiencing bothersome PMS symptoms.

**Trial registration:** NCT07013539, date of registration 02/06/2025

## Background

The global prevalence of premenstrual syndrome (PMS) is high affecting nearly 50% of women of reproductive age worldwide^1^. PMS is a significant public health burden in modern societies due to its impact on women’s quality of life^2^, contributing to reduced work productivity, increased absenteeism, or increased risk of suicidal ideation leading to a substantial economic burden on both individuals and society^3,4^.

According to the American College of Obstetricians and Gynecologists, symptoms of PMS should be present during the 5 days before a period for at least three menstrual cycles in a row, end within 4 days after a period starts and interfere with some normal activities^5^. These symptoms follow a cyclical pattern and are categorized into psychological, physical, and behavioral groups. The most common reported symptoms include irritability, anxiety, mood swings, agitation, anger, insomnia, difficulty concentrating, lethargy, depression, and severe fatigue. Fluid retention may cause edema, transient weight gain, breast fullness and pain. Pelvic heaviness or pressure and backache may also occur^6–8^.

Despite the high prevalence and burden of PMS, its pathophysiology remains incompletely understood. Consequently, there is no pharmacologic treatment specific to PMS. Current evidence suggests that PMS is not associated with abnormal ovarian hormone concentrations, as estrogen and progesterone levels are generally comparable between affected and unaffected women ^9^. Rather, it has been suggested that susceptible women may exhibit an abnormal central nervous system (CNS) response to the normal cyclical fluctuations of ovarian steroids during the luteal phase associated with dysfunction of the serotonin and γ-aminobutyric acid (GABA) neurotransmitter systems^5^. Indeed, estrogen has been shown to modulate serotonergic neurotransmission by regulating serotonin synthesis, metabolism, transporter activity, and receptor expression. Consequently, cyclical fluctuations in ovarian hormones may transiently disrupt serotonergic signaling in susceptible women, contributing to the mood and behavioral symptoms characteristic of PMS^9,10^. This hypothesis is supported by symptom exacerbation following acute tryptophan depletion, the primary precursor of serotonin, as well as by the clinical efficacy of selective serotonin reuptake inhibitors (SSRIs), which remain the first-line pharmacological treatment for the management of affective premenstrual disorders^5^. However, SSRIs are frequently associated with adverse events, limiting their long-term acceptability and highlighting the need for safer therapeutic alternatives. Beyond its role in mood regulation, serotonin also plays an important role in descending pain modulation pathways, suggesting that altered serotonergic neurotransmission may contribute not only to the emotional manifestations of PMS but also to its physical symptomatology^10,11^.

Increasing evidence also highlights the importance of γ-aminobutyric acid (GABA) neurotransmission^12^. The progesterone metabolite, allopregnanolone, is a potent positive allosteric modulator of GABA-A receptors and normally exerts anxiolytic effects during the luteal phase. In women with PMS or PMDD, altered sensitivity of GABA-A receptors to physiological fluctuations in allopregnanolone appears to impair inhibitory neurotransmission, resulting in increased neuronal excitability and greater vulnerability to anxiety, irritability, mood instability, and emotional dysregulation.

However, fluctuations in ovarian steroids may also influence the renin–angiotensin–aldosterone system (RAAS), vasopressin secretion, and microvascular permeability, leading to transient sodium and water retention^13,14^. In parallel, progesterone-related changes in gastrointestinal motility and visceral sensitivity may contribute to abdominal bloating and the sensation of pelvic heaviness, while low-grade inflammation may further exacerbate tissue edema and discomfort^15^.

In addition to neuroendocrine mechanisms, accumulating evidence suggests that low-grade inflammation, oxidative stress, and dysregulation of the hypothalamic-pituitary-adrenal (HPA) axis may contribute to symptom development through complex interactions with neurotransmitter systems and neuroactive steroids^16^. Increased circulating inflammatory cytokines (interleukines IL-1, 4, 7, 8 and 10) ^17^ and impaired antioxidant status have been reported in women with moderate-to-severe PMS, although findings remain heterogeneous^2,17^. Inflammatory mediators may also increase endothelial permeability and contribute to fluid extravasation, providing a potential mechanistic link with edema and abdominal discomfort^18^. Altogether, these observations support the concept that PMS is a multifactorial disorder involving hormonal sensitivity, neurotransmitter dysfunction, oxidative stress, inflammatory processes, altered fluid homeostasis and psychosocial factors.

Given this multifactorial pathophysiology, therapeutic strategies capable of simultaneously targeting serotonergic dysfunction, GABAergic signaling, oxidative stress, and inflammation are of increasing interest. Thanks to its bioactive compounds, safranal and crocins, known for its antioxidant, anti-inflammatory, and its modulatory role on serotonin neurotransmission^19–23^, saffron extract could be a natural and safer solution to improve women’s health including PMS symptoms^24^.

To our knowledge, few randomized, double-blind, placebo-controlled trials conducted in Iranian women suffering from PMS suggested promising results of saffron extract^25^. Agha-Hosseini and al. demonstrated that 15mg saffron extract administered twice a day in 50 women aged 20-45 with PMS significantly improved premenstrual and depressive symptoms within two menstrual cycles in comparison to a placebo, as assessed with the Total Premenstrual Daily Symptoms and the Hamilton rating scale^26^. Similarly, Pirdadeh Beiranvand and al. showed that 30mg of dried extract of saffron stigma once a day during two menstrual cycles significantly reduced the severity of PMS symptoms over time, in 88 university students suffering from this syndrome^27^. Although the two trials suggest that saffron extract may reduce PMS severity, the current evidence remains preliminary and has important methodological and external-validity limitations. These include modest sample sizes, short treatment duration, unstandardized saffron extract, limited statistical methods by current standards, and restriction to Iranian populations, including a highly selected student sample. Consequently, a new, adequately powered, well-controlled clinical trial in Western women who experience different socio-economic and cultural conditions is warranted to confirm efficacy and strengthen generalizability. This double-blind, randomized, placebo-controlled, clinical trial addresses this gap by investigating the effect of standardized saffron extract in women suffering from PMS during 3 menstrual cycles of supplementation.

## Materials and methods

### Study design

This randomized, multicentric, double-blind, parallel-group, placebo-controlled trial was conducted from September 2025 to June 2026 at five study sites from Complife Italia S.r.l: San Martino Siccomario, Biella, Garbagnate Milanese, Milan and Rende. All study procedures were conducted in accordance with the ethical principles outlined in the World Medical Association’s (WMA) Declaration of Helsinki and its amendments. The study protocol and Informed Consent Form (ICF) were reviewed by “Comitato Etico Indipendente per le Indagini Cliniche Non Farmacologiche” (Genova, Italy) and approved on 29 May 2025 (Rif. 2025/12). The clinical trial was registered on clinicaltrial.gov (NCT07013539). No important changes to methods were made after trial commencement. All participants provided written informed consent before any study procedures.

### Participants

One hundred and ten healthy Caucasian women, aged between 18 and 45 years, were enrolled in the study according to the following inclusion criteria: mild-to-moderate PMS symptoms for at least three menstrual cycles, regular menstrual cycles (21–35 days, menstrual flow 2–8 days), and body mass index between 18.5 and 24.9 kg/m². PMS eligibility was prospectively assessed by a psychologist over three consecutive cycles using the Premenstrual Symptoms Screening Tool (PSST), a reliable and validated self-assessment questionnaire designed to screen women experiencing premenstrual syndrome^28^ and menstrual regularity was documented using a menstrual diary in which participants indicated the duration of their menstrual cycles and the duration of their menstrual flow.

Exclusion criteria were perimenopause, acute or chronic conditions potentially interfering with the study such as polycystic ovary syndrome, endometriosis, thyroid disease, chronic fatigue syndrome, irritable bowel syndrome, and psychiatric disorders including depression, anxiety, and bipolar disorder, pregnancy or breastfeeding, food allergy or intolerance to study ingredients, recent use of similar food supplements, and use of medications such as antidepressants, anxiolytics, diuretics, hormonal contraceptives, and non-steroidal anti-inflammatory drugs.

### Randomization and blinding

Participants were randomized in a 1:1 ratio to receive either the saffron extract or placebo. The randomization list was generated by an independent technician from Complife Italia S.r.l, not involved in the enrollment, using the Wey’s urn algorithm implemented in PASS 11 software (version 11.0.8; PASS, LLC, Kaysville, UT, USA) and stored securely. The study was conducted under double-blind conditions: participants, investigators, and all the study staff were unaware of study product allocation until the end of statistical analyses. The treatments were provided by Activ’Inside in opaque and sealed bottles and were indistinguishable in appearance, packaging, and labeling, differing solely by batch numbers.

### Treatments and compliance

Participants were randomly allocated to receive one bottle of 95 hard-shell capsules, each containing 30 mg of saffron extract (Safr’Inside^TM^, Activ’Inside, Beychac et Caillau, France, obtained through the patent EP3490575B1-WO2017EP69200 containing crocins *>*3 % and safranal *>*0.2 %, analyzed using the U-HPLC-DAD method) or maltodextrin (placebo). At baseline, participants were instructed to take one capsule of study product per day, during breakfast with a glass of water, starting supplementation approximately 7 days before their next menstruation and to continue daily intake until the end-of-treatment visit after three menstrual cycles. Treatment compliance was assessed by study staff using participant treatment diaries, and by counting the numbers of remaining capsules in the bottles at each visit. Compliance with treatment was calculated as follows:

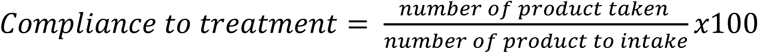

Participants were considered compliant if intake ranged between 80% and 120% of the prescribed regimen over the 3-month period.

### Study flow

After database pre-screening, women attended a first screening visit (T−4) at which eligibility was reviewed. They were informed about the study procedures and were asked to sign an informed consent form and an authorization for the processing of personal data. Demographic data were collected, and participants received the PSST and menstrual diary. During the subsequent three cycles (T−3, T−2, T−1), the PSST was completed monthly and reviewed by a psychologist to confirm mild-to-moderate PMS.

At the baseline visit (T0), scheduled 7 ± 2 days after the start of menstrual flow, participants returned their menstrual diary, eligibility was reconfirmed, and they completed the Premenstrual Assessment Form–Short Form (PAF-SF) and the Patient Health Questionnaire-4 (PHQ-4). Participants were then randomized and received a bottle containing saffron or placebo capsules and a treatment diary.

Follow-up visits were performed 7 ± 2 days after the onset of menstrual flow, allowing for the assessment of both PMS symptoms and menstrual symptoms. At T1, this corresponded to approximately 7 days of supplementation during the PMS phase and 14 ± 2 days of supplementation by the time of the follow-up visit. As subsequent visits were scheduled according to menstrual cycle length, T2 and T3 occurred within broader planned windows and corresponded approximately to 35 and 63 days of supplementation, respectively. At each visit, menstrual diaries were collected, eligibility was reviewed, and PAF-SF and PHQ-4 questionnaires were completed. At T3, participants returned unused capsules, and treatment compliance was assessed by capsule count and treatment diary review. A final menstrual diary was returned at T4, 1–2 days after the start of the next menstrual flow.

### Outcomes

#### Premenstrual Assessment Form - Short Form

The PAF-SF is a validated, reliable, self-administered questionnaire used to assess the severity of PMS symptoms^29^. It consists of 10 items evaluating changes in psychological and physical symptoms on a 6-point Likert scale ranging from 1 (“no change”) to 6 (“extreme change”). Item scores are summed to generate a total PAF-SF score, which was defined as the primary study outcome, as well as three subscale scores reflecting affect, water retention, and pain. The total score ranges from 10 to 60, with higher scores indicating greater symptom severity. Although the PSST is a validated and clinically useful instrument for screening women with PMS^28^, the PAF-SF was considered more appropriate as the primary endpoint because it offers a concise, continuous, symptom-severity score that is better suited to detecting longitudinal changes in response to supplementation ^29^.

#### Patient Health Questionnaire-4

The PHQ-4 is a validated, reliable, self-administered screening questionnaire used to assess core symptoms of anxiety and depression^30^. It consists of 4 items rated on a 4-point Likert scale ranging from 0 (“not at all”) to 3 (“nearly every day”). Item scores are summed to derive a total PHQ-4 score, as well as two 2-item subscale scores: the anxiety and the depression subscales. The total score ranges from 0 to 12, with higher scores indicating greater psychological distress. Each subscale score ranges from 0 to 6, with higher scores indicating greater severity of anxiety or depressive symptoms, respectively.

#### Adverse events

The tolerability of the product was closely monitored by the study staff throughout the study period. Subjects had access to the investigator via a contact phone number provided with the study information sheet in case of adverse reactions. If a subject reported an event, the study staff established whether it is related to the study product.

#### Sample size

To our knowledge, only two published studies have evaluated the effects of 30 mg saffron supplementation on PMS symptoms in Iranian women aged less than 50 years^26,27^. Among these, only one provided sufficiently detailed numerical data by treatment group and time point to support sample size estimation. In that study, a significant time-by-group interaction was reported, indicating that changes in mean PMS severity differed significantly between groups over time (p < 0.001). Based on these data, the sample size was calculated using PROC GLMPOWER in SAS software for a repeated-measures ANOVA with three PMS score assessments. Although the initial sample size calculation indicated that 40 participants would be sufficient for the primary endpoint analysis, 110 women were recruited to enhance the robustness of the study. This larger sample size was chosen to account for the expected variability of PMS symptoms across individuals and menstrual cycles, to mitigate the impact of dropouts or incomplete follow-up, and to improve the precision and external validity of the findings.

#### Statistical analysis

The primary analysis was conducted on the intention-to-treat (ITT) population, defined as all randomised participants with at least one available post-randomisation measurement. A per-protocol (PP) analysis was performed as a sensitivity analysis and included all participants who completed the study without major protocol deviations; participants were excluded from the PP population if they missed one or more evaluation visits or did not use the study product as instructed during the intervention period. Baseline demographic and clinical characteristics were summarized using the mean and standard deviation (SD).

Efficacy endpoints collected at four timepoints (T0, T1, T2 and T3) were analysed using linear mixed-effects models with restricted maximum likelihood (REML) estimation. For each endpoint, the model included treatment group, time, and treatment-by-time interaction as fixed effects, together with a subject-specific random intercept to account for within-subject correlation across repeated measurements. In the primary confirmatory analysis, time was modelled as a continuous variable; therefore, the treatment-by-time interaction represented the between-group difference in the rate of change over time and constituted the main test of treatment effect. The primary endpoint was the PAF-SF total score. P-values were derived using the Satterthwaite approximation for degrees of freedom, and 95% confidence intervals were calculated using the Wald method.

To identify the timepoint at which between-group differences emerged, a secondary supportive analysis was performed using a mixed-effects model of the same structure but with time specified as a categorical variable (T0, T1, T2 and T3). This allowed treatment differences to be estimated separately at each post-baseline visit on change from baseline, following a difference-in-differences approach that is not affected by baseline imbalance between groups. Because these visit-specific comparisons involved multiple post-baseline tests, they were considered exploratory and were adjusted within each outcome using the Holm procedure across T1, T2 and T3.

All statistical tests were two-sided, and statistical significance was set at p<0.05. Analyses were performed using R software (version 4.2.1; R Core Team), with the lme4 and lmerTest packages for mixed-effects modelling and emmeans for estimated marginal means and multiplicity-adjusted comparisons.

## Results

A total of 110 women were randomised, with 55 assigned to the saffron group and 55 to placebo. The ITT population included all randomised participants with at least one available post-randomisation assessment (55 per group). The PP population comprised 54 participants in the saffron and 53 in the placebo group. Three participants discontinued the study for personal reasons unrelated to the study product: one in the saffron group and two in the placebo group. Overall, 107 participants completed the study (54 in the active group and 53 in the placebo group). Details of participant baseline demographic and clinical information are detailed in Table 1.

**Table 1.** Baseline characteristics of the population.

|  | Total<br>n=110<br>Mean (SD) | Placebo<br>n=55<br>Mean (SD) | Saffron<br>extract<br>n=55<br>Mean (SD) |
| --- | --- | --- | --- |
| **Age (years)** | 30.5 (8.0) | 31.7 (8.3) | 29.3 (7.6) |
| **Body mass index (kg/m<sup>2</sup>)** | 21.7 (1.8) | 21.6 (1.8) | 21.9 (1.7) |
| **Duration of menstrual cycle (days)** | 28.5 (2.5) | 28.4 (2.0) | 28.5 (2.9) |
| **Duration of menstruations (days)** | 5.3 (0.9) | 5.2 (0.9) | 5.4 (1.0) |
| **Premenstrual Assessment Form Short Form** |  |  |  |
| Total score | 32.4 (7.3) | 31.2 (7.1) | 33.6 (7.2) |
| Affect score | 13.4 (3.3) | 13.2 (2.1) | 13.6 (3.4) |
| Pain score | 9.7 (2.4) | 9.3 (2.4) | 10.2 (2.4) |
| Water retention score | 9.3 (2.9) | 8.7 (3.0) | 9.9 (2.7) |
| **Patient Health Questionnaire-4** |  |  |  |
| Total score | 4.5 (1.7) | 4.6 (1.6) | 4.5 (1.8) |
| Anxiety score | 2.6 (1.0) | 2.6 (0.9) | 2.6 (1.1) |
| Depression score | 1.9 (1.1) | 2.0 (1.2) | 1.9 (1.1) |

The mean age of participants was 31±8 years and they have a mean body mass index of 22±2 kg/m². Women had a baseline menstrual cycle of 28±3 days on average and menstrual flow of 5±1 days on average, in accordance with inclusion criteria. The mean PAF-SF total score of 32±7 and a mean PHQ-4 score of 4.5±2 indicated moderate premenstrual symptoms and a low level of anxiety and depressive symptoms, respectively. During the intervention phase, menstrual cycle length and menstrual flow duration remained consistent across cycles and were comparable between treatment groups (data not shown).

### Primary endpoint: PAF-SF Total score

The PAF-SF total score decreased over time in both groups, with a greater reduction in the saffron group than in the placebo group. In the primary analysis (Table 2), the treatment-by-time interaction was statistically significant (difference in slope: -1.88; 95% CI -2.64 to -1.12; p<0.001), indicating a significantly greater reduction in total PAF-SF score over time in the saffron group than in the placebo group. In the supportive visit-specific analysis of change from baseline (Figure 1), the between-group difference was significant from T1 onward and increased over time: -2.42 at T1 (95% CI -4.79 to -0.04; adjusted p=0.046), -3.40 at T2 (95% CI -5.78 to -1.02; adjusted p=0.010), and -5.96 at T3 (95% CI -8.34 to -3.58; adjusted p<0.001). The PP analysis yielded materially identical results (data not shown).

**Figure 1.**
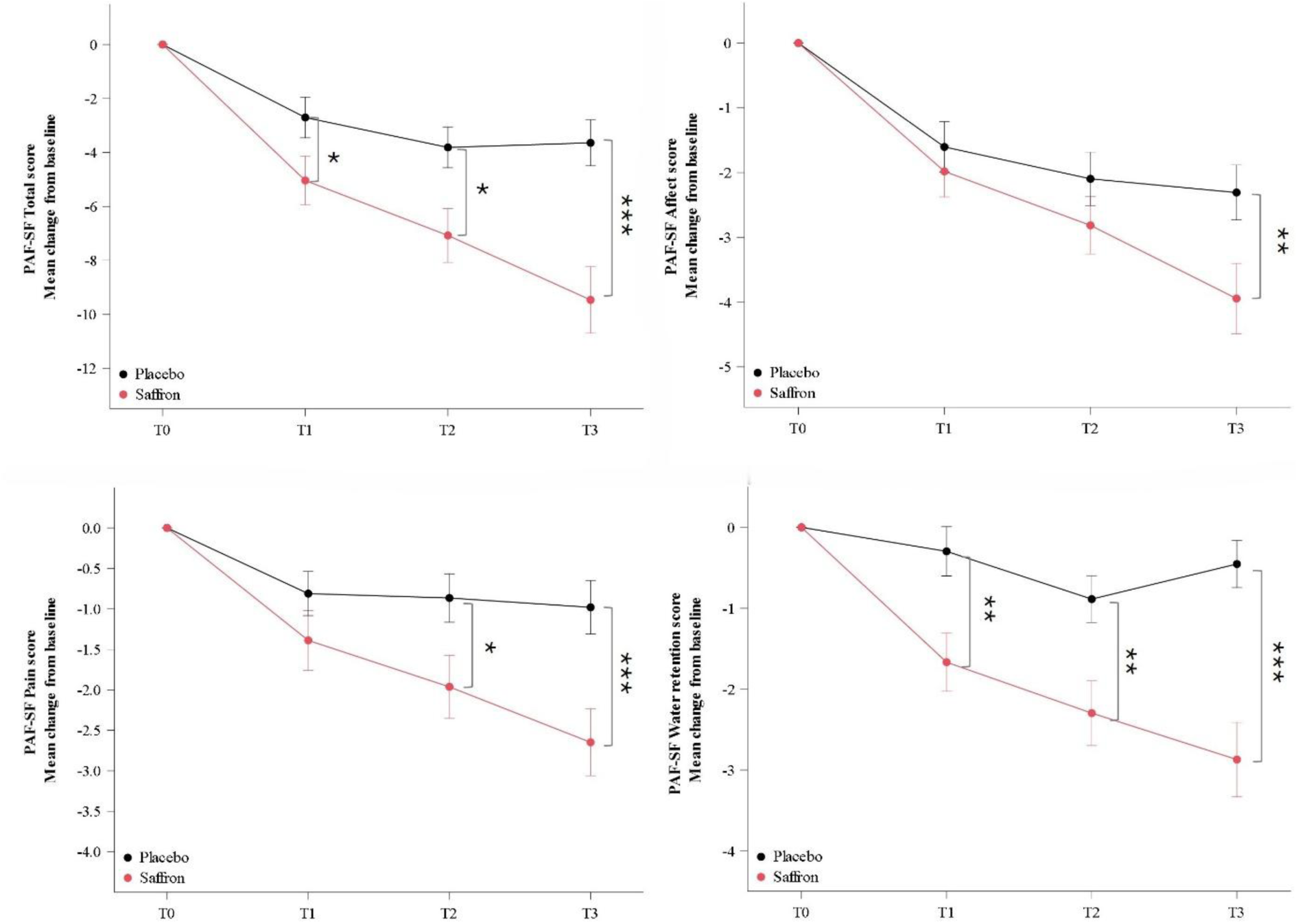
Changes in PAF-SF scores across the study period (mean ± SE). T1, first menstrual cycle after 7 days of supplementation; T2, second menstrual cycle after 35 days of supplementation; T3, third menstrual cycle after 63 days of supplementation. Holm-adjusted p-values for post-hoc between-group comparisons were derived from linear mixed-effects models: *p < 0.05, **p < 0.01, ***p < 0.001.

**Table 2.**
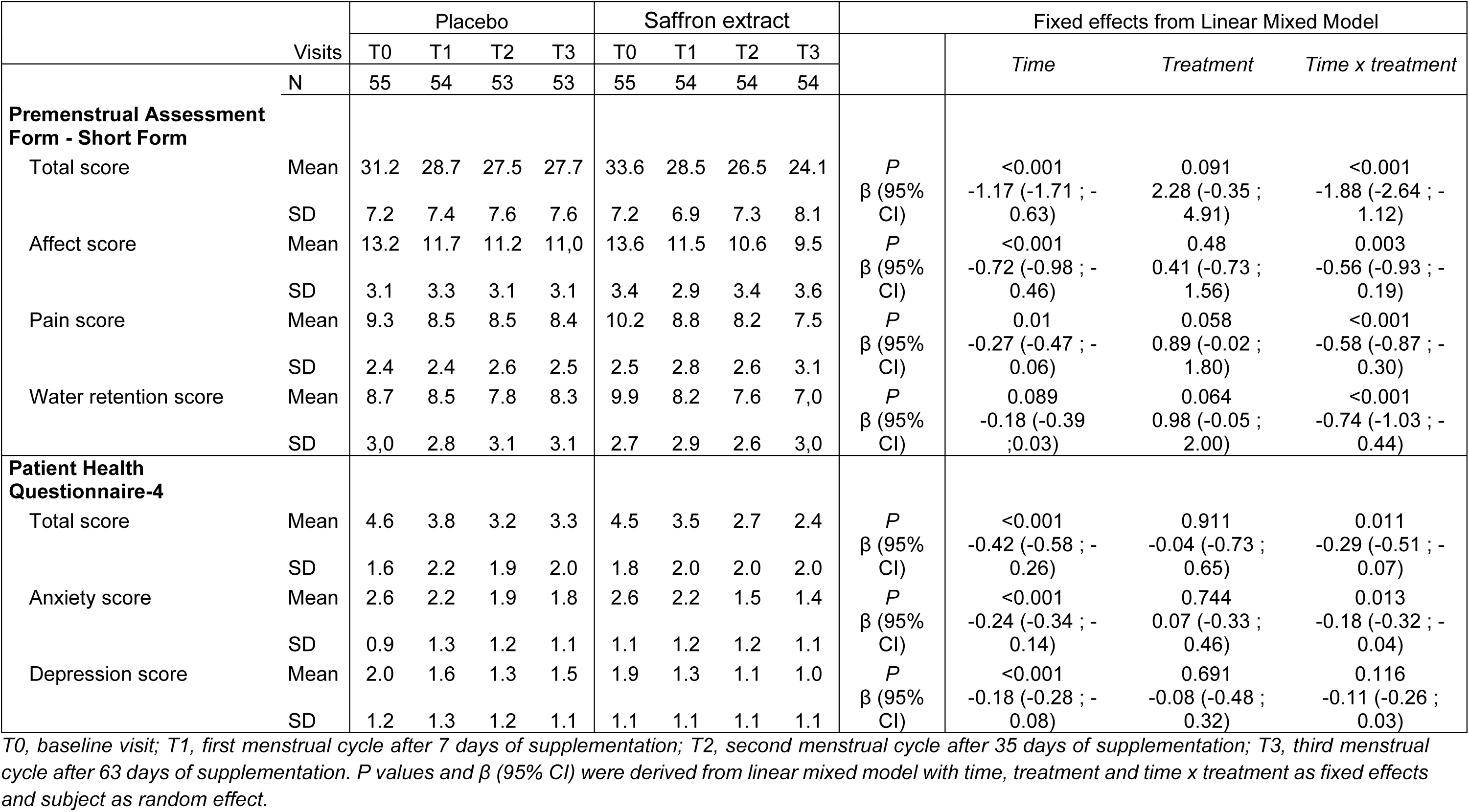
Effect of saffron extract supplementation on premenstrual and psychological symptoms over 3 menstrual cycles (N=110)

### Secondary endpoints

#### Affect

The PAF-SF affect subscale also improved in both groups, with a larger decrease in the saffron group. The treatment-by-time interaction was statistically significant (difference in slope: -0.56; 95% CI -0.93 to -0.19; p=0.003), supporting a greater improvement in affective symptoms in the saffron group. In visit-specific analyses, the between-group difference in change from baseline was not significant at T1 or T2 but became significant at T3 (-1.76; 95% CI -2.92 to -0.61; adjusted p=0.009).

#### Pain

The PAF-SF pain subscale showed progressive improvement over time in both groups, with greater improvement in the active group. The treatment-by-time interaction was significant (difference in slope: -0.58; 95% CI -0.87 to -0.30; p<0.001), indicating a significantly greater reduction in pain symptoms in the saffron group. In visit-specific analyses, the difference between-group change from baseline was not significant at T1 but became significant at T2 (-1.15; 95% CI -2.05 to -0.24; adjusted p=0.026) and T3 (-1.76; 95% CI -2.66 to -0.85; adjusted p<0.001).

#### Water retention

A distinct pattern was observed for the PAF-SF water retention subscale. Although the overall time effect was not statistically significant in the pooled analysis (p=0.089), the treatment-by-time interaction was highly significant (difference in slope: -0.74; 95% CI -1.03 to -0.44; p<0.001), indicating a more favorable trajectory in the saffron group. In visit-specific analyses, the between-group difference in change from baseline was already significant at T1 (-1.38; 95% CI -2.31 to -0.46; adjusted p=0.0048), remained significant at T2 (-1.44; 95% CI -2.37 to -0.51; adjusted p=0.005), and further increased at T3 (-2.45; 95% CI -3.37 to -1.52; adjusted p<0.001).

#### PHQ-4

The PHQ-4 total score decreased modestly in both groups, with greater improvement in the saffron group. The mixed-effects model showed a significant treatment-by-time interaction (difference in slope: -0.29; 95% CI -0.51 to -0.07; p=0.011), indicating a significantly greater improvement in the saffron group over time. In visit-specific analyses, the between-group difference in change from baseline became significant at T3 only (-0.85; 95% CI -1.55 to -0.16; adjusted p=0.0498).

Similarly, the anxiety score decreased in both groups, with greater improvement in the saffron group. The mixed-effects model showed a significant treatment-by-time interaction (difference in slope: -0.18; 95% CI -0.32 to -0.04; p=0.013), indicating a significantly greater improvement in the saffron group over time. However, the between-group difference in change from baseline in visit-specific analyses were not statistically significant.

The depression score significantly decreased over time in both groups, as showed by the time effect (p < 0.001) but the pattern of change over time was not significantly different between the two groups (time x treatment; p = 0.116). PP analyses yielded to similar results for all secondary endpoints: all time x treatment p-values remained statistically significant (p<0.05, data not shown).

#### Compliance

Treatment compliance was excellent in both groups. All participants were considered compliant. The mean compliance was 100% in each intervention group.

#### Adverse events

No adverse events were documented during the study period. The three recorded discontinuations were all due to personal reasons and were considered unrelated to the study product. Overall, the study product was well-tolerated.

## Discussion

To our knowledge, this is the first randomized, double-blind, placebo-controlled trial conducted in Western women to evaluate the effects of a standardized saffron extract on PMS symptoms over three menstrual cycles. In women with mild-to-moderate PMS, daily supplementation with 30 mg saffron extract resulted in a significantly greater reduction in overall PMS symptom severity than placebo. This benefit emerged from the first menstrual cycle and increased progressively across the third cycle. Improvements were also observed across the main symptom domains captured by the PAF-SF—affect, pain and water retention—as well as on the PHQ-4 measure of psychological distress. Taken together, these findings support the potential of Safr’Inside^TM^ as a well-tolerated non-pharmacological option for women experiencing bothersome PMS symptoms.

Beyond demonstrating a significant reduction in symptom scores, these findings provide additional insight into the multidimensional nature of PMS^5^. PMS is increasingly recognized not as a single entity but as a heterogeneous syndrome resulting from interactions between ovarian steroid fluctuations, CNS sensitivity, neurotransmitter regulation, immune signaling, and peripheral physiological responses. Therefore, the observation that this standardized saffron extract influenced several symptom domains suggests that its potential effects may extend beyond a single pathway and may involve multiple biological processes contributing to PMS symptom generation.

An important contribution of the present study is that it extends the existing evidence base beyond the small number of prior randomized controlled trials conducted in Iranian women. The two existing clinical trials reported that saffron supplementation improved PMS symptom severity and depressive symptoms compared with placebo after two menstrual cycles [^26,27^]. In the earlier trial by Agha-Hosseini et al., 15 mg saffron extract administered twice daily significantly reduced Total Premenstrual Daily Symptoms scores relative to placebo ^26^. Similarly, Pirdadeh Beiranvand et al. found that 30 mg/day saffron reduced PMS severity over time in university students^27^. Our results are consistent with these previous findings and strengthen the evidence supporting a beneficial effect of saffron on PMS symptoms. At the same time, the current study substantially improves the external validity of the literature by including Western adult women recruited in a multicenter setting, rather than a selected student sample. This is particularly relevant because PMS expression, symptom perception, stress exposure, diet, and help-seeking behaviors may vary across cultural and socioeconomic contexts [7,8]. Demonstrating efficacy in this context therefore adds important information regarding the generalizability of previous findings.

The overall symptom improvement observed with saffron is biologically plausible considering current understanding of PMS pathophysiology. PMS is no longer considered a consequence of abnormal ovarian hormone concentrations per se, but rather of an increased sensitivity of the CNS to normal cyclical fluctuations in gonadal steroids ^31,32^. This sensitivity appears to involve altered serotonergic and GABAergic signaling, as well as abnormal responses to neuroactive steroid metabolites such as allopregnanolone ^31,33,34^. The established efficacy of SSRIs in PMS and PMDD strongly supports the role of serotonin in symptom generation and modulation ^5,35^. In this context, the present findings are coherent with preclinical and clinical evidence indicating that saffron and its bioactive constituents may modulate monoaminergic neurotransmission, stress-related pathways, and neuroinflammatory processes^20–23^.

In our study, the PAF-SF affect subscale and the PHQ-4 total score improved significantly more in the saffron group over time, with the between-group difference for PHQ-4 becoming statistically significant after three menstrual cycles of supplementation. These findings are consistent with prior evidence suggesting beneficial effects of saffron on symptoms of anxiety, depression, and stress^36–38^. Several meta-analyses and randomized trials outside the PMS field have reported antidepressant or anxiolytic effects of saffron comparable in magnitude to standard pharmacological treatments in mild-to-moderate mood disorders^39–41^, although these bodies of evidence are heterogeneous in terms of populations, formulations, and methodological quality. The fact that the PHQ-4 effect emerged later than the reduction in total PAF-SF score may reflect the relatively low baseline level of anxiety and depressive symptoms in our sample, leaving less room for change on this instrument than on the PMS-specific questionnaire.

The improvement in pain-related symptoms is also noteworthy. Pain is a major component of PMS and may include headache, backache, musculoskeletal pain, abdominal discomfort, and breast tenderness^6,7^. In our study, the pain subscale improved significantly more with saffron than with placebo, with between-group differences apparent from T2 onward. Although the underlying mechanisms remain incompletely understood, this finding is biologically plausible given the reported anti-inflammatory and antinociceptive properties of saffron and its main constituents, crocins, crocetin and safranal^42,43^. Supporting this hypothesis, Pourtau et al. reported that circulating metabolites derived from Safr’Inside^TM^ intake modulated pro-inflammatory and pro-catabolic mediators in human cellular models under inflammatory conditions, suggesting a potential systemic anti-inflammatory activity^44^. Preclinical evidence further suggests that saffron may modulate nociceptive signaling through multiple complementary mechanisms ^45^. In particular, its constituents have demonstrated anti-inflammatory and antioxidant activities that may attenuate the inflammatory and oxidative processes potentially contributing to nociceptor sensitization. In addition, safranal has been shown to modulate the Transient Receptor Potential A1 (TRPA1) ion channel expressed by nociceptive neurons, leading to subsequent channel desensitization and reduced Calcitonine Gene-Related Peptide (CGRP) release and nociceptive responses^46^. Such effects could contribute to limiting nociceptor sensitization and pain signaling and may therefore, at least partly, account for the improvement in premenstrual pain observed in the present study. However, these mechanisms remain hypothetical in the context of PMS and warrant confirmation in clinical studies^44^. Clinical findings provide further indirect support for a potential analgesic effect of saffron, as beneficial effects have been reported in menstrual-related pain conditions such as dysmenorrhea^25,47^. Overall, these findings suggest that the effects of saffron in PMS may involve not only emotional regulation but also modulation of inflammatory and pain-related pathways.

The findings observed for the water retention domain are also of particular interest. The treatment-by-time interaction was highly significant, with between-group differences becoming apparent as early as T1 and increasing over time. Water retention related symptoms—including bloating, swelling, and transient weight gain—are common and often highly bothersome manifestations of PMS and contribute substantially to its overall burden^7,31^. Unlike affective symptoms, these physical symptoms are thought to relate more directly to changes in fluid and sodium homeostasis, vascular regulation, and gastrointestinal function^7,47^. During the luteal phase, progesterone acts as a physiological antagonist of the mineralocorticoid receptor, triggering compensatory activation of the RAA system. The subsequent decline in progesterone before menstruation may allow aldosterone-mediated sodium and water retention to become more pronounced, thereby contributing to bloating, edema, and breast tenderness ^13,14^. Low-grade inflammation may also exacerbate tissue edema and physical discomfort in susceptible women ^18^. Although no evidence currently indicates that saffron directly modulates the RAA system, its anti-inflammatory, antioxidant, and potentially vasoactive properties, together with preclinical evidence suggesting diuretic activity, may contribute to reduce symptom severity or the perception of fluid retention^44,48–50^. This early effect may also have contributed to the improvement in total PAF-SF score. Future mechanistic studies should explore whether saffron influences biological correlates with bloating and fluid retention or other pathways potentially involved in these symptoms.

As expected, the tolerability profile observed in this study was favorable. No adverse events related to the study product were documented, and compliance was excellent in both groups. This is clinically relevant because currently recommended pharmacological options for PMS disorders, especially SSRIs, are effective but may be limited by side effects such as nausea, insomnia, somnolence, fatigue, and impaired concentration. A safe and acceptable nutritional intervention may therefore be of particular interest for women with mild-to-moderate PMS who seek symptom relief but are reluctant to use medication or do not require prescription treatment.

This study has several strengths. It used a randomized, double-blind, placebo-controlled design, was conducted across multiple centers, included a relatively large sample (n=110) compared with previous PMS saffron trials, and assessed symptoms repeatedly throughout the screening and intervention periods. Eligibility was prospectively documented over three cycles using the PSST, a validated screening tool, together with a menstrual diary, which increases confidence that participants genuinely experienced recurrent PMS rather than nonspecific distress. The use of both ITT and PP analyses, which yielded materially similar findings, also supports the robustness of the results. Furthermore, the study evaluated both PMS-specific outcomes and broader psychological distress, allowing a more nuanced interpretation of treatment effects.

Some limitations should nonetheless be acknowledged. First, this population was restricted to healthy Western women with mild-to-moderate PMS, normal BMI, and regular menstrual cycles, limiting generalizability to women with severe PMS or PMDD, adolescents, or perimenopausal women. Second, although three cycles represent a meaningful duration for PMS research, longer follow-up would be valuable to determine the persistence of benefit and to assess whether saffron efficacy plateaus, increases, or wanes over time. Third, while the PSST was used prospectively for eligibility assessment, the primary endpoint was the PAF-SF because its items provide a more detailed assessment of physical symptoms and allow the derivation of affective, pain, and water-retention subscores. Fourth, the study was not powered for formal subgroup analyses, such as differential response according to baseline symptom profile, age, or psychological distress level. Finally, multiplicity remains a consideration for secondary and visit-specific analyses; although exploratory comparisons were adjusted using the Holm procedure, these analyses should be interpreted as supportive rather than definitive.

## Conclusion

In this randomized, double-blind, placebo-controlled trial conducted in Western women with mild-to-moderate PMS, daily supplementation with 30 mg Safr’Inside^TM^ for three menstrual cycles resulted in a significantly greater improvement in overall PMS symptom severity than placebo. This effect was evident from 7 days post-supplementation during the first menstrual cycle and increased progressively over the course of supplementation. Specifically, significant benefits were observed in the main symptom domains of PMS, including affective, pain, and water retention symptoms. The favorable tolerability profile and excellent treatment compliance further support the potential of saffron extract as a well-tolerated and acceptable non-pharmacological approach for women seeking relief from bothersome PMS symptoms. Further clinical studies are warranted to confirm these results in broader populations, including women with more severe PMS, and to investigate the long-term persistence of benefits.

## Data Availability

All data produced in the present study are available upon reasonable request to the authors

## List of abbreviations

BMI: Body mass index
CGRP: Calcitonine Gene-Related Peptide
GABA: Gamma-aminobutyric acid
ITT: Intention-to-threat
PAF-SF: Premenstrual Assessment Form - Short Form
PHQ-4: Patient Health Questionnaire 4
PMS: Premenstrual syndrome
PP: Per-protocol
PSST: Premenstrual syndrome screening tool
RAAS: Renine-Angiotensine-Aldosterone system
SD: Standard deviation
SE: Standard error of the mean
SSRIs: Selective serotonin reuptake inhibitors
TRPA: Transient Receptor Potential A1
Tx: Timepoint x

## Declarations

## Ethics approval and consent to participate

The study was conducted in accordance with the Declaration of Helsinki and approved (ref. 2025/12) by the “Independent Ethical Committee for Non-Pharmacological Clinical trials” (Genova, Italy) on 24/02/2026. Informed consent was obtained from all subjects involved in the study.

## Consent for publication

Not applicable.

## Availability of data and materials

The raw data supporting the conclusions of this article will be made available by the authors on request. The data are not publicly available since they are the property of the sponsor of the study (Activ’Inside, Beychac et Caillau, France).

## Funding

This research and the APC was funded by Activ’Inside (grant R209836) who received support for the Silver Brain Food project, a program co-financed by the “Future Investment Program” (Programme d’Investissements d’Avenir PIA3) and managed by the Investment General Secretariat and operated by Bpifrance.

### Acknowledgements

The authors would like to express their gratitude to the Complife Italia staff, who contributed to the study and recruited the subjects, for their professionalism and support during study development.

